# Parental smoking in children consulting for respiratory diseases in Switzerland

**DOI:** 10.64898/2026.08.17.26360586

**Authors:** Tayisiya Krasnova, Maša Žarković, Carina Nigg, Mari Sasaki, Mandukhai Ganbat, Carmen Casaulta, Alexander Moeller, Nicolas Regamey, Claudia E Kuehni, the SPAC Study Team

**Affiliations:** Institute of Social and Preventive Medicine, University of Bern, Bern, Switzerland; Graduate School for Health Sciences, University of Bern, Bern, Switzerland; Division of Paediatric Respiratory Medicine and Allergology, Department of Paediatrics, Inselspital, Bern University Hospital, University of Bern, Switzerland; Department of Respiratory Medicine, University Children’s Hospital Zurich and Children’s Research Center, University of Zurich, Zurich, Switzerland; Division of Paediatric Pulmonology, Children’s Hospital of Central Switzerland, Lucerne, Switzerland

**Keywords:** Children, tobacco smoke, lung diseases, respiratory outpatient clinics, surveys and questionnaires

## Abstract

**Background:** Exposure to environmental tobacco smoke (ETS) negatively affects children’s health, but few studies examined parental smoking behaviour in families of children with respiratory diseases. We studied parental smoking prevalence, characteristics, and changes over one year among families in the Swiss Paediatric Airway Cohort (SPAC).

**Methods:** We included children aged 0–17 years referred to paediatric respiratory outpatient clinics in Switzerland from 2017 to 2024. Parents answered a questionnaire at the initial clinic visit and again after one year. We used multivariable logistic regression to explore the characteristics of mothers and fathers who smoked and assessed changes in smoking behavior over one year.

**Results:** Among 4,199 children (median age 9 years [IQR 5–12]), 31% were exposed to parental smoking at baseline (paternal smoking: 16%; maternal smoking: 6%; both parents smoking: 9%). Mothers were more likely to smoke if they had a lower education level (OR 2.0, 95%CI 1.6–2.5 for compulsory education vs university education), did not have Swiss nationality (OR 1.3, 1.0–1.6) and lived in a socially disadvantaged neighborhood (OR 1.3, 1.0–1.7). Similar associations were observed for fathers. In addition, fathers were more likely to smoke if they were unemployed (OR 2.0, 1.3–3.2 vs having a full-time job. The strongest predictor of smoking was having a partner who smoked, with ORs above 6 for both mothers and fathers.

Parents of 2,338 children completed the one-year follow-up questionnaire. Data from 2226 mothers and 1895 fathers showed that among baseline smokers with follow-up data, 225 (78%) mothers and 382 (81%) of fathers continued smoking, and only 63 (22%) of mothers and 90 (19%) of fathers quit. Among baseline non-smokers, 47 (2%) mothers and 54 (3%) fathers started smoking.

**Conclusions:** One-third of children consulting respiratory specialists in Switzerland are exposed to parental smoking. ETS exposure was strongly associated with socio-economic factors. Even after visiting a specialized clinic, most parents continued to smoke. This highlights the urgent need for stronger national smoking policies and targeted support to help these parents quit and stay smoke-free.

## Introduction

Exposure to environmental tobacco smoke (ETS) is detrimental for children’s health ^1–3^. It impairs mucociliary clearance, weakens immune function, increases susceptibility to respiratory infections and risk of allergic sensitisation, and can trigger inflammation and chronic cough in children ^4–6^. For children already experiencing respiratory symptoms, ETS exposure increases the frequency and severity of symptoms, leading to more hospital visits and poorer overall health ^7–11^. Parental smoking is particularly harmful as children spend much time with their parents and at home. Additionally, it strongly influences children’s likelihood of adopting the habit themselves, perpetuating a harmful cycle ^12–16^. Addressing parental smoking is key to protecting children’s health. Globally, approximately 40% of children are exposed to ETS ^2^. The prevalence of smoking remains high in Switzerland, with 55% of men and 42% of women being current or former smokers in 2022 ^17^. Around one-third of children in Switzerland grow up in households where at least one parent smokes ^18,19^. We know less about smoking in families with children who have a respiratory disease. Understanding how common parental smoking is and which families are most affected is crucial for designing effective strategies to reduce children’s exposure to tobacco smoke. In this study we investigated parental smoking in children referred to specialized clinics due to respiratory symptoms in Switzerland. We assessed the prevalence of parental smoking at the time of the clinical visit and one year later, and explored factors associated with maternal and paternal self-reported tobacco smoking and potential changes in smoking behavior over one year.

## Methods

### The Swiss Paediatric Airway Cohort (SPAC)

The Swiss Paediatric Airway Cohort (SPAC) is a national, prospective, multicenter study established in 2017 ^20^. It enrolls patients aged 0-17 years, referred to respiratory outpatient clinics or pulmonology practices for common symptoms such as cough, wheezing, or exercise-induced breathing difficulties, excluding those with severe pre-existing conditions like cystic fibrosis or cardiac disorders. The study is observational, integrated into routine care, and follows local protocols.

SPAC collects data at several timepoints: initially through the baseline questionnaire completed at the first clinical visit and then through annual follow-up questionnaires. In addition, the SPAC study team collects data from patients’ medical records, including referral letters and outpatient clinic letters with information on suspected or confirmed diagnoses, results of diagnostic tests, and prescribed treatments (Figure S1). Ethical approval was granted by the Bern Cantonal Ethics Committee (KEK 2016–02176), and informed consent is obtained from parents and children aged 14 or older.

### Study population and inclusion criteria

We included SPAC participants from respiratory outpatient clinics in Aarau, Basel, Bern, Chur, Lausanne, Lucerne, St. Gallen, Zurich and pulmonology practices in Worb and Horgen between July 2017 and July 2024.

### Smoking outcomes

In the baseline questionnaire, participants reported on respiratory symptoms, personal and family medical history, environmental exposures, and sociodemographic factors.

The questionnaire specifically asked, “Does the mother/father smoke in the household?”, separately for mothers and fathers. We coded current parental smoking as a binary variable (yes/no), where “yes” indicated that the parent responded “Yes, also indoors” or “Yes, only outdoors” and “no” indicated “No, never smoked” or “No, not anymore”.

The follow-up questionnaire one year after the clinical visit contained the same smoking-related questions, except that for paternal smoking, “Father or other person” was offered instead of just “Father.” To remain consistent between questionnaires, responses indicating father, partner, or stepfather were classified as paternal smoking, whereas responses indicating any other person were treated as missing for paternal smoking.

### Explanatory variables

We assessed sociodemographic characteristics separately for each parent, guided by previous literature. These included Swiss language regions (German- or French-speaking); nationality (Swiss or non-Swiss for all other nationalities); education level, categorized as primary (compulsory schooling), secondary (apprenticeship, vocational school, commercial or technical school), or tertiary (teacher training seminars, higher technical college at least 3 years, or university); and employment status, grouped as full-time job, part-time job, multiple part-time jobs, or unemployed/in education. We additionally included age of the child, categorized into 0–5, 6–11, or 12–17 years; the child’s sex (male or female); and the number of children in the family (1, 2 or >2). We also included the Swiss Neighborhood Index of Socioeconomic Position (Swiss-SEP), which ranges from 0 to 100, with higher values indicating a higher socioeconomic position ^21^. We assigned Swiss-SEP values to the geographical coordinates of the children’s home address and categorized values into high, medium, or low tertiles, following Panczak et al ^25^. We also classified the household location by Urban/Rural Typology 2020 into three categories (urban, periurban and rural) based on the municipalities according to the classification of the Federal Statistical Office ^22^.

### Statistical analysis

We used medians and interquartile ranges (IQR) to summarize age, and percentages with 95% confidence interval (95%CI) to describe categorical variables. To explore associations between parental smoking at the first clinical visit and sociodemographic characteristics, we conducted a multivariable logistic regression. Initially, we fitted univariable logistic regression models to identify sociodemographic factors associated with maternal and paternal smoking. Variables with a p-value <0.1 were included in the multivariable model. We a priori included language region, which has known differences in Swiss smoking rates ^17^, and child’s sex, which might be associated with exposure to parental smoking for girls and boys ^23^, in the final model. We excluded household location from the model because of its strong correlation with the Swiss-SEP. As a sensitivity analysis, we additionally fitted models including questionnaire year as a covariate.

We compared changes in parental smoking between baseline and the one-year follow-up using the McNemar test for paired data among participants with complete information. To account for potential bias due to differential loss to follow-up, inverse probability weights were calculated from logistic regression models estimating the probability of participation at follow-up based on baseline parental smoking and sociodemographic characteristics. Stabilized weights were applied to obtain weighted estimates of smoking prevalence and transitions, representing the baseline population. All analyses were performed using Stata (Version 16.1, Stata Corporation, Austin, TX).

## Results

### Characteristics of the study population

By July 2024 4,199 families had completed the baseline questionnaire. Median age of the child was 8.5 years [IQR 5–12]; 60% were male (Table 1, Figure S2, Sample A). Most (94%) lived in the German-speaking region, 16% had one child, 55% had two, and 27% had more than two children. Twenty-nine percent of families were classified as living in low, 33% in medium, and 38% in high socioeconomic neighborhoods, respectively. Sixty percent lived in urban areas, 24% in peri-urban, and 16% in rural. Most participating mothers (75%) and fathers (74%) reported Swiss nationality. Secondary education was the most frequently attained education level among mothers (47%), while fathers more often attained tertiary education (45%). Employment status differed between parents with 8% of mothers, and 79% of fathers reporting a full-time job (Table 1, Table S3).

**Table 1.**
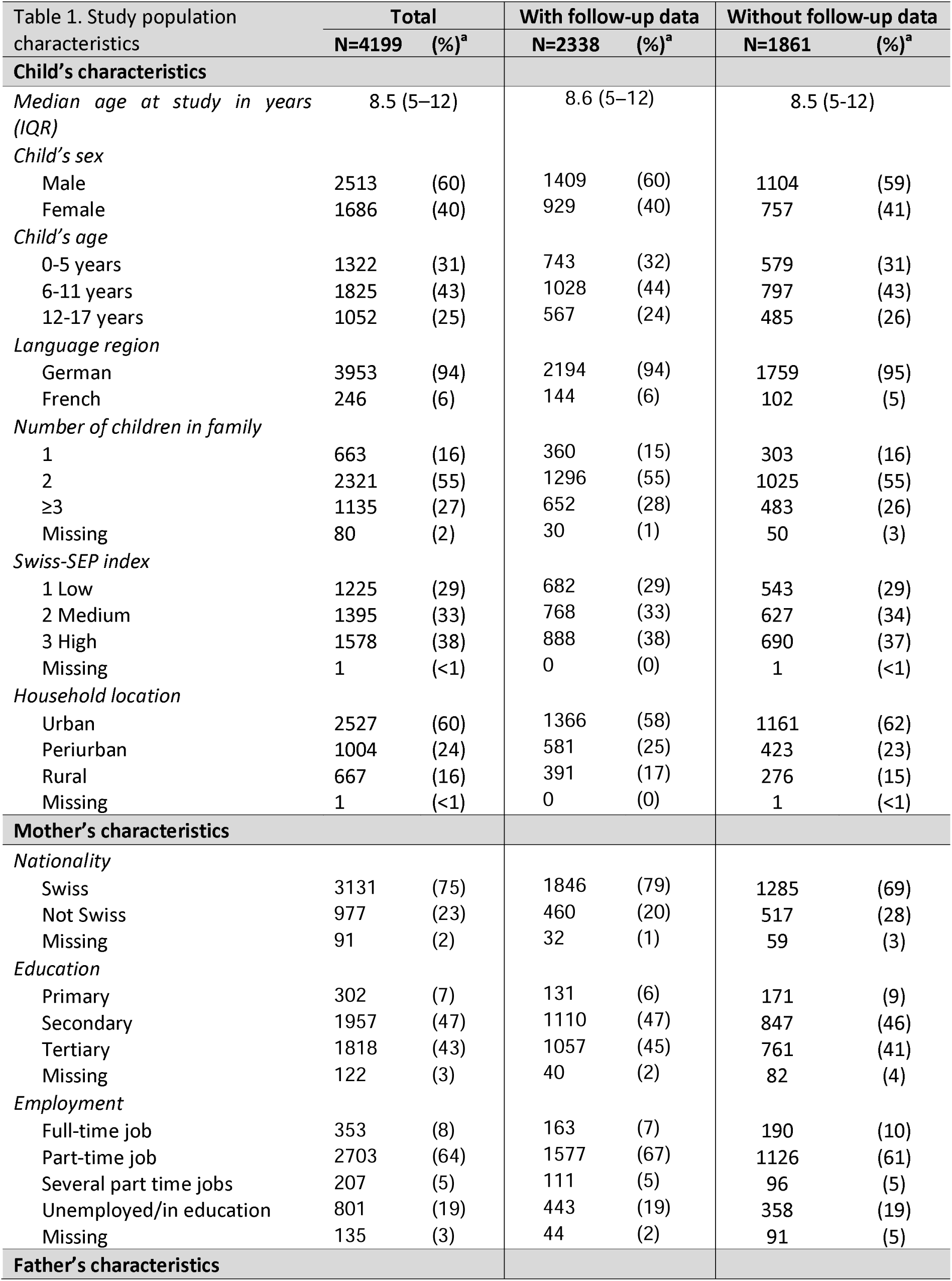
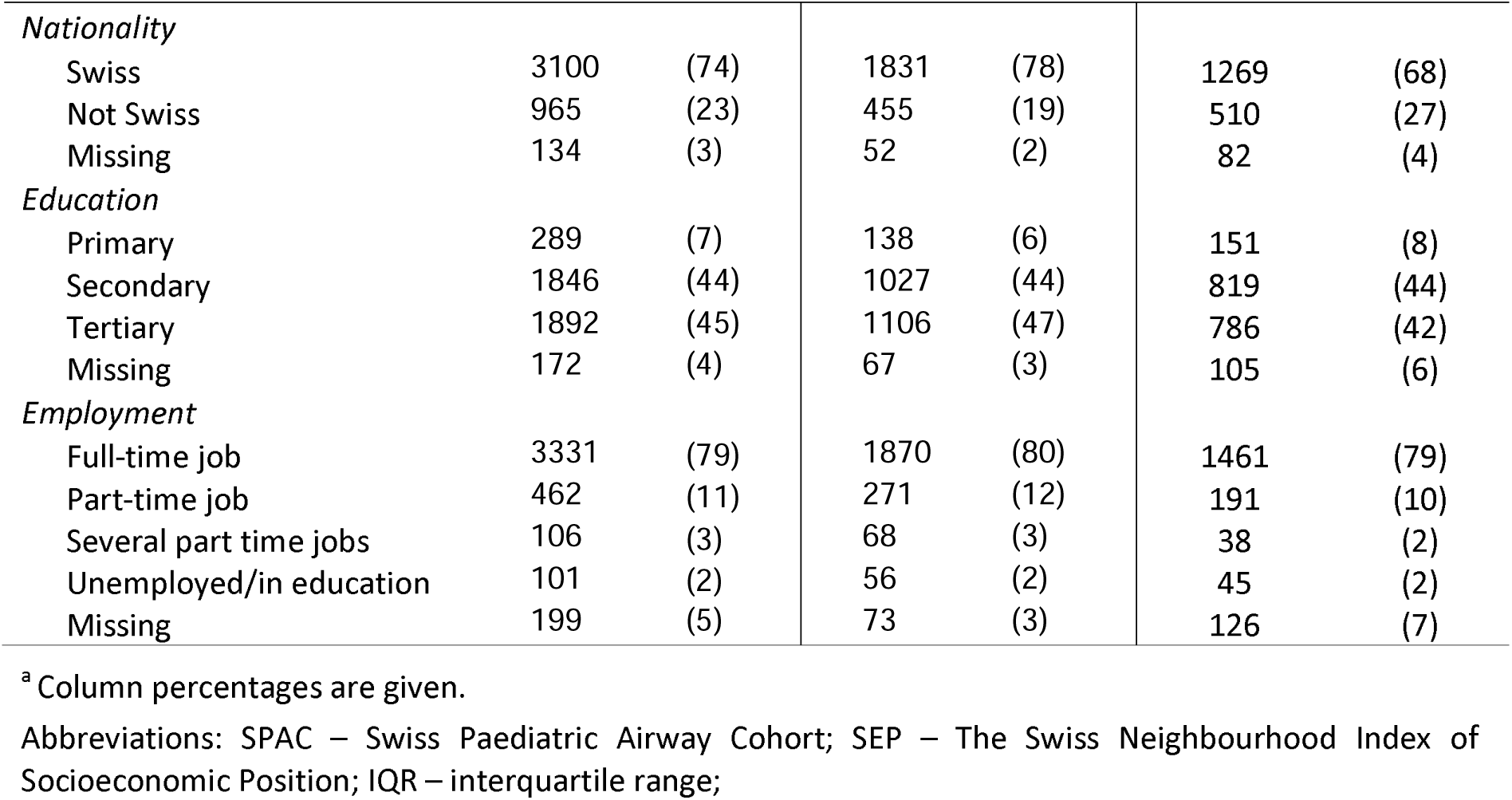
Characteristics of participants of the Swiss Paediatric Airway Cohort, comparing those with and without follow-up data.

| Table 1. Study population characteristics | Total<br>N=4199 (%)<sup>a</sup> |  | With follow-up data<br>N=2338 (%)<sup>a</sup> |  | Without follow-up data<br>N=1861 (%)<sup>a</sup> |  |
| --- | --- | --- | --- | --- | --- | --- |
| **Child's characteristics** |  |  |  |  |  |  |
| *Median age at study in years (IQR)* | 8.5 (5–12) |  | 8.6 (5–12) |  | 8.5 (5–12) |  |
| *Child's sex* |  |  |  |  |  |  |
| Male | 2513 | (60) | 1409 | (60) | 1104 | (59) |
| Female | 1686 | (40) | 929 | (40) | 757 | (41) |
| *Child's age* |  |  |  |  |  |  |
| 0-5 years | 1322 | (31) | 743 | (32) | 579 | (31) |
| 6-11 years | 1825 | (43) | 1028 | (44) | 797 | (43) |
| 12-17 years | 1052 | (25) | 567 | (24) | 485 | (26) |
| *Language region* |  |  |  |  |  |  |
| German | 3953 | (94) | 2194 | (94) | 1759 | (95) |
| French | 246 | (6) | 144 | (6) | 102 | (5) |
| *Number of children in family* |  |  |  |  |  |  |
| 1 | 663 | (16) | 360 | (15) | 303 | (16) |
| 2 | 2321 | (55) | 1296 | (55) | 1025 | (55) |
| ≥3 | 1135 | (27) | 652 | (28) | 483 | (26) |
| Missing | 80 | (2) | 30 | (1) | 50 | (3) |
| *Swiss-SEP index* |  |  |  |  |  |  |
| 1 Low | 1225 | (29) | 682 | (29) | 543 | (29) |
| 2 Medium | 1395 | (33) | 768 | (33) | 627 | (34) |
| 3 High | 1578 | (38) | 888 | (38) | 690 | (37) |
| Missing | 1 | (<1) | 0 | (0) | 1 | (<1) |
| *Household location* |  |  |  |  |  |  |
| Urban | 2527 | (60) | 1366 | (58) | 1161 | (62) |
| Periurban | 1004 | (24) | 581 | (25) | 423 | (23) |
| Rural | 667 | (16) | 391 | (17) | 276 | (15) |
| Missing | 1 | (<1) | 0 | (0) | 1 | (<1) |
| **Mother's characteristics** |  |  |  |  |  |  |
| *Nationality* |  |  |  |  |  |  |
| Swiss | 3131 | (75) | 1846 | (79) | 1285 | (69) |
| Not Swiss | 977 | (23) | 460 | (20) | 517 | (28) |
| Missing | 91 | (2) | 32 | (1) | 59 | (3) |
| *Education* |  |  |  |  |  |  |
| Primary | 302 | (7) | 131 | (6) | 171 | (9) |
| Secondary | 1957 | (47) | 1110 | (47) | 847 | (46) |
| Tertiary | 1818 | (43) | 1057 | (45) | 761 | (41) |
| Missing | 122 | (3) | 40 | (2) | 82 | (4) |
| *Employment* |  |  |  |  |  |  |
| Full-time job | 353 | (8) | 163 | (7) | 190 | (10) |
| Part-time job | 2703 | (64) | 1577 | (67) | 1126 | (61) |
| Several part time jobs | 207 | (5) | 111 | (5) | 96 | (5) |
| Unemployed/in education | 801 | (19) | 443 | (19) | 358 | (19) |
| Missing | 135 | (3) | 44 | (2) | 91 | (5) |
| **Father's characteristics** |  |  |  |  |  |  |
| *Nationality* |  |  |  |  |  |  |
| Swiss | 3100 | (74) | 1831 | (78) | 1269 | (68) |
| Not Swiss | 965 | (23) | 455 | (19) | 510 | (27) |
| Missing | 134 | (3) | 52 | (2) | 82 | (4) |
| *Education* |  |  |  |  |  |  |
| Primary | 289 | (7) | 138 | (6) | 151 | (8) |
| Secondary | 1846 | (44) | 1027 | (44) | 819 | (44) |
| Tertiary | 1892 | (45) | 1106 | (47) | 786 | (42) |
| Missing | 172 | (4) | 67 | (3) | 105 | (6) |
| *Employment* |  |  |  |  |  |  |
| Full-time job | 3331 | (79) | 1870 | (80) | 1461 | (79) |
| Part-time job | 462 | (11) | 271 | (12) | 191 | (10) |
| Several part time jobs | 106 | (3) | 68 | (3) | 38 | (2) |
| Unemployed/in education | 101 | (2) | 56 | (2) | 45 | (2) |
| Missing | 199 | (5) | 73 | (3) | 126 | (7) |

<sup>a</sup> Column percentages are given.
Abbreviations: SPAC – Swiss Paediatric Airway Cohort; SEP – The Swiss Neighbourhood Index of Socioeconomic Position; IQR – interquartile range;

At the time of the analysis, 3,698 families had been enrolled in the cohort for more than 12 months and had therefore received a follow-up questionnaire. A total of 2,338 families (63%) returned the questionnaire. Compared to non-responders, responders to the follow-up questionnaire were more often Swiss citizens (80% vs. 70–71%), had higher education (46% tertiary vs. 39% primary education for mothers; 49% vs. 41% for fathers), and were more likely to have a non-smoking partner (77% vs. 68% for mothers; 87% vs. 79% for fathers). Among mothers, those who responded were more frequently employed part-time (69% vs. 62%) and were less often unemployed (19% vs. 22%). (Table S2).

### Prevalence of parental smoking

At enrolment, 1,290 out of 4199 (31%, 95% CI 31-34%) children were exposed to parental smoking (Table 2). Among these, 651 (16%, 95%CI 15-17%) were exposed exclusively to paternal smoking, 249 (6%, 95%CI 5-7%) to maternal smoking, and 390 (9%, 95%CI 9-11%) to both parents’ smoking. Regarding smoking intensity, 10% of mothers and 13% of fathers reported to smoke ≤ 10 cigarettes per day, while 4% of mothers and 11% of fathers smoked >10 cigarettes daily. At the one-year follow-up, 144 of 2226 mothers (7%, 95%CI 6-8%) and 315 of 1895 fathers (16%, 95%CI 14-17%) were smokers. In 133 families (7%, 95%CI 6-8%), both parents were smokers.

**Table 2.** Prevalence of parental smoking in participants of the Swiss Paediatric Airway Cohort at the first visit and one year later.

| Any smoking | Baseline |  | One-year follow-up |  |
| --- | --- | --- | --- | --- |
|  | N=4199 | (%)<sup>a</sup> | N=2338 | (%)<sup>a</sup> |
| None | 2724 | (65) | 1428 | (61) |
| Maternal only | 249 | (6) | 144 | (6) |
| Paternal only | 651 | (16) | 315 | (13) |
| Both parents | 390 | (9) | 133 | (6) |
| Missing | 185 | (4) | 318 | (14) |
| **Maternal smoking** | **N=4199** | **(%)<sup>a</sup>** | **N=2226** | **(%)<sup>a</sup>** |
| Smoker | 639 | (15) | 272 | (12) |
| Non-smoker | 3478 | (83) | 1954 | (88) |
| Missing | 82 | (2) | 0 | (0) |
| *Cigarettes number per day* |  |  |  |  |
| Non-smoker | 3478 | (83) | 1954 | (88) |
| ≤10 | 438 | (10) | 193 | (9) |
| >10 | 164 | (4) | 66 | (3) |
| Missing | 119 | (3) | 13 | (1) |
| **Paternal smoking** | **N=4199** | **(%)<sup>a</sup>** | **N=1895** | **(%)<sup>a</sup>** |
| Smoker | 1041 | (25) | 436 | (23) |
| Non-smoker | 2955 | (70) | 1459 | (77) |
| Missing | 203 | (5) | 0 | (0) |
| *Cigarettes number per day* |  |  |  |  |
| Non-smoker | 2955 | (70) | 1459 | (77) |
| ≤10 | 525 | (13) | 253 | (13) |
| >10 | 456 | (11) | 162 | (9) |
| Missing | 263 | (6) | 21 | (1) |

<sup>a</sup> Column percentages are given.
Abbreviations: SPAC – Swiss Paediatric Airway Cohort;

### Associations between maternal smoking and sociodemographic factors

Compared to non-smoking mothers, mothers who smoked had lower education (primary education: OR = 1.7, 95% CI 1.1–2.5; secondary education: OR = 2.0, 95% CI 1.6–2.6, compared to tertiary education), more often had non-Swiss nationality (OR = 1.3, 95% CI 1.0–1.6), lived in socioeconomically disadvantaged neighborhoods (OR = 1.3, 95% CI 1.0–1.7), and had a partner who smokes (OR = 6.9, 95% CI 5.6–8.3). Additionally, mothers who smoked tended to have fewer children (one child: OR = 1.6, 95% CI 1.2– 2.2; two children: OR = 1.3, 95% CI 1.0–1.6 compared to 3 and more children) (Table 3, Figure S3).

**Table 3.** Associations of maternal and paternal smoking with sociodemographic characteristics at the baseline clinical visit in the Swiss Paediatric Airway Cohort, from two multivariable logistic regression models.

|  | Maternal smoking<br>N = 3759<sup>b</sup> |  |  | Paternal smoking<br>N = 3696<sup>b</sup> |  |  |
| --- | --- | --- | --- | --- | --- | --- |
|  | OR | (95% CI) | p<sup>a</sup> | OR | (95% CI) | p<sup>a</sup> |
| *Child's sex* |  |  | **0.023** |  |  | 0.231 |
| Male | Ref. |  |  | Ref. |  |  |
| Female | 1.3 | (1.0-1.5) |  | 0.9 | (0.8-1.0) |  |
| *Child's age* |  |  | 0.770 |  |  | **0.037** |
| 12-17 years | Ref. |  |  | Ref. |  |  |
| 6-11 years | 1.0 | (0.8-1.3) |  | 1.0 | (0.8-1.2) |  |
| 0-5 years | 1.1 | (0.8-1.4) |  | 1.3 | (1.0-1.6) |  |
| *Language region* |  |  | 0.290 |  |  | 0.134 |
| German | Ref. |  |  | Ref. |  |  |
| French | 1.2 | (0.8-1.9) |  | 0.8 | (0.5-1.1) |  |
| *Number of children in family* |  |  | **0.012** |  |  | 0.943 |
| ≥3 | Ref. |  |  | Ref. |  |  |
| 2 | 1.3 | (1.0-1.6) |  | 1.0 | (0.8-1.2) |  |
| 1 | 1.6 | (1.2-2.2) |  | 1.0 | (0.8-1.3) |  |
| *Swiss-SEP index* |  |  | 0.064 |  |  | **0.040** |
| 3 High | Ref. |  |  | Ref. |  |  |
| 2 Medium | 1.3 | (1.0-1.6) |  | 1.1 | (0.9-1.3) |  |
| 1 Low | 1.3 | (1.0-1.7) |  | 1.3 | (1.1-1.6) |  |
| *Nationality* |  |  | 0.072 |  |  | **<0.001** |
| Swiss | Ref. |  |  | Ref. |  |  |
| Not Swiss | 1.3 | (1.0-1.6) |  | 1.5 | (1.2-1.8) |  |
| *Education* |  |  | **<0.001** |  |  | **<0.001** |
| Tertiary | Ref. |  |  | Ref. |  |  |
| Secondary | 2.0 | (1.6-2.6) |  | 2.3 | (1.9-2.7) |  |
| Primary | 1.7 | (1.1-2.5) |  | 2.2 | (1.6-3.1) |  |
| *Employment* |  |  | 0.138 |  |  | **0.012** |
| Full-time job | Ref. |  |  | Ref. |  |  |
| Part-time job | 0.8 | (0.5-1.1) |  | 1.1 | (0.8-1.4) |  |
| Several part time jobs | 1.1 | (0.6-1.8) |  | 1.5 | (0.9-2.5) |  |
| Unemployed/in education | 1.0 | (0.7-1.4) |  | 2.0 | (1.3-3.2) |  |
| *Partner who smokes* |  |  | **<0.001** |  |  | **<0.001** |
| Non-smoker | Ref. |  |  | Ref. |  |  |
| Smoker | 6.9 | (5.6-8.3) |  | 6.4 | (5.2-7.8) |  |

<sup>a</sup> P – P-value calculated from likelihood-ratio test. Ref.—Reference
<sup>b</sup> N reflects respondents with complete data on all variables included in the model
Abbreviations: OR – Odds Ratio; CI – Confidence Interval; SEP – Swiss Neighbourhood Index of Socioeconomic Position;

**Figure 1.**
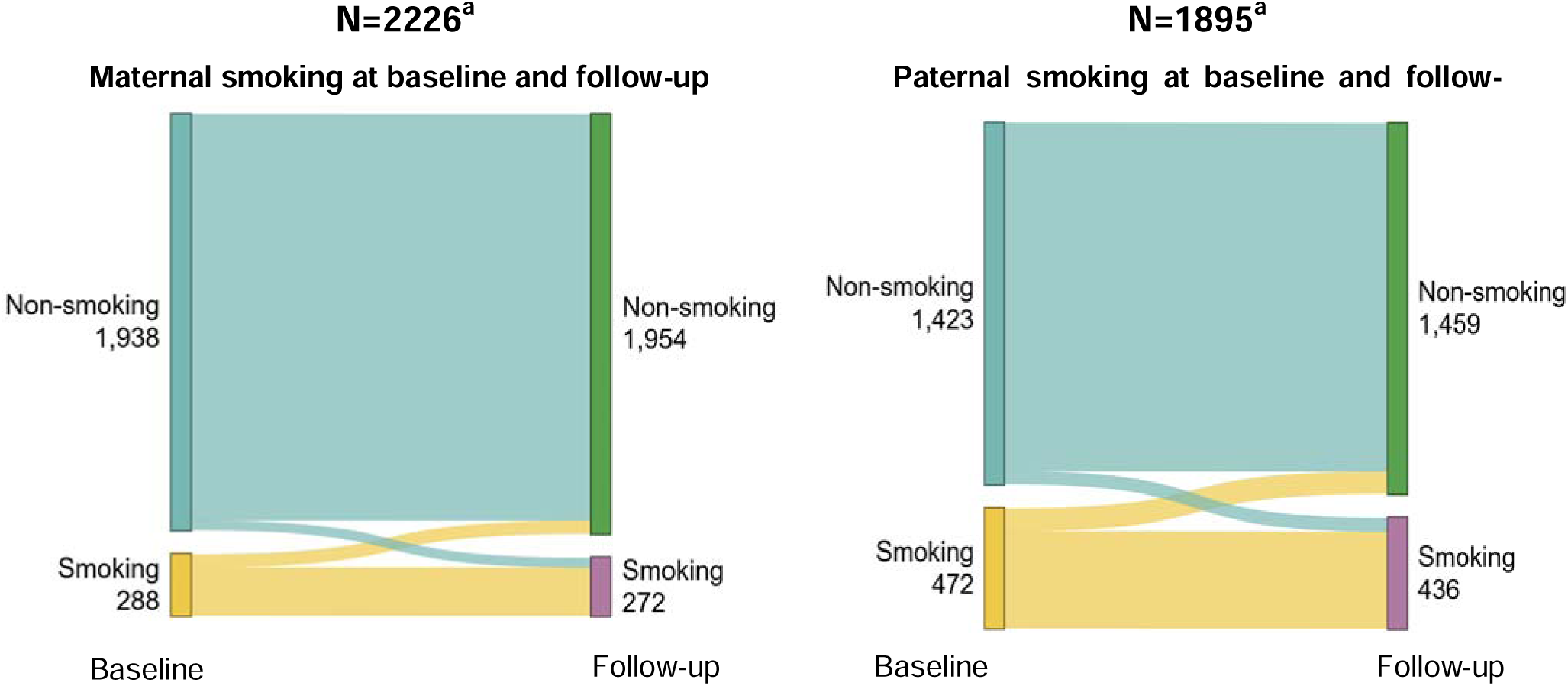
Changes in maternal and paternal smoking status from baseline to one-year follow-up in the Swiss Paediatric Airway Cohort, illustrated using alluvial plots. Yellow indicates smokers and blue indicates non-smokers. ^a^ N reflects respondents with complete smoking data at both baseline and 1-year follow-up questionnaires.

### Associations between paternal smoking and sociodemographic factors

Compared to non-smoking fathers, fathers who smoked had lower education (primary education: OR = 2.2, 95% CI 1.6–3.1; secondary education: OR = 2.3, 95% CI 1.9–2.7, compared to tertiary education), more often had non-Swiss nationality (OR = 1.5, 95% CI 1.2–1.8), lived in socioeconomically disadvantaged neighborhoods (OR = 1.3, 95% CI 1.1–1.6), more often had a smoking partner (OR = 6.4, 95% CI 5.2–7.8). However, unlike mothers, fathers who smoked were more likely to be unemployed (OR = 2.0, 95% CI 1.3–3.2, compared to full-time employment) and had younger children (0–5 years: OR = 1.3, 95% CI 1.0–1.6, compared to children aged 12–17 years) (Table 3, Figure S4). Results were unchanged in sensitivity analyses additionally adjusting for questionnaire year in the maternal and paternal models.

### Changes in smoking behavior over one year

Among the 2,338 families who returned the follow-up questionnaire, 2,226 answered the question on maternal smoking status and 1,895 on paternal smoking status (Figure S2, *Sample B and Sample C*). Over the one-year period, 225 (10%) mothers and 382 (20%) fathers continued smoking, 47 mothers (2%) and 54 (3%) fathers started smoking, while only 63 mothers (3%) and 90 fathers (4%) had quit. Of those who started smoking over the one-year period, 32 (68%) mothers and 35 (65%) fathers were ex-smokers at baseline.

Inverse probability weighting indicated that smokers and parents with lower socioeconomic status were less likely to participate at follow-up, modestly increasing estimated smoking prevalence among mothers (12% to 14%) and fathers (23% to 25%). Baseline smoking remained a strong predictor of continued smoking, as did having a partner who smokes.

## Discussion

This study found that one third of children referred to respiratory specialists in Switzerland are exposed to parental tobacco smoking. Both maternal and paternal smoking were strongly associated with socio-economic factors, and the majority of parents continued to smoke after one year, showing little effect of the visit to the specialist centre.

Smoking prevalence among parents in our cohort (15% of mothers and 25% of fathers) was lower than in the general Swiss population reported in the Swiss Health Survey 2017 and 2022 (23% of women in 2017 and 21% in 2022 and 31% of men in 2017 and 27% in 2022) ^17,19^ and in population-based studies from Zurich in 2013-2016 (20% and 30%) ^13^, the SCARPOL study in 1999 (39% and 34%) ^7^, and the HBSC survey in 2010 (41%) ^24^. This difference may partly reflect the overall decline in cigarette smoking rates in Switzerland since 1992 ^25^ and potentially greater health awareness among families of children with respiratory disease. However, underreporting due to social desirability is also possible, particularly in parents of children with respiratory problems, as a survey among the Swiss residents in 2012-2015 showed that smoking prevalence is up to 45% lower than estimates from tobacco sales ^26, 27^. If present, such bias would lead to an underestimation of parental smoking in our study.

Parents with lower education, non-Swiss nationality, living in disadvantaged neighbourhoods, and with a smoking partner were more likely to smoke, in line with previous Swiss studies^7,17,19^. These findings confirm that socioeconomic determinants are an important driver of parental smoking. Limited health literacy, psychosocial stress, and targeted tobacco marketing may contribute to these disparities^28,29^, while the lack of comprehensive tobacco control policies in Switzerland^30^ may further perpetuate them^31^.

Previous reports^17,19,28^ showed higher smoking rates among employed parents. In our study smoking was more common among unemployed fathers and did not differ by employment among mothers. This may be due to differences in families with children and gendered employment patterns in Switzerland, where women are less likely to work full-time^32,33^, and where unemployment-related stress may increase smoking among fathers^34,35^. The 2022 Tobacco Consumption report found higher smoking in the French-speaking region^17^, which we did not observe, perhaps due to the small number of participants from this region in our study. And notably, we lack data on parental age, a factor known to influence smoking behavior due to its strong association with declining tobacco consumption with aging ^37–41^. Thus, the observed associations between parental smoking, child’s age, and family size may be partially explained by parental age.

Smoking behaviour changed little over one year: most smokers continued, few quit, and a similar number initiated smoking. Notably, about two-thirds of new smokers were former smokers, highlighting relapse as a major issue and identifying recent quitters as a particularly vulnerable group ^36^. In participating outpatient clinics, parents are routinely asked about smoking and, if positive, advised to quit and referred to cessation programs, but no standardized intervention has been implemented. Our findings suggest that this procedure has little measurable effect. If parental smoking in this high-risk population is to be reduced, more efficient and easily accessible smoking cessation programs, ideally free of charge and embedded within tertiary care centres, are needed. Even more importantly, stronger tobacco control policies in Switzerland that make smoking more difficult and more expensive are likely essential to achieve population-level impact.

### Strengths and limitations

Our study focuses on a particularly vulnerable population — children with respiratory diseases, who are more susceptible to harmful effects of passive smoking ^7–11^, providing insights into this at-risk group in Switzerland. The multi-center clinics large sample size makes it more representative for Switzerland. Loss to follow-up may have introduced selection bias, but weighted analyses suggest only modest effects on prevalence estimates.

## Conclusion

This study conducted in tertiary respiratory outpatient clinic in Switzerland showed that most parents continued smoking at the follow-up visit, indicating that specialist consultations alone have no smoking cessation effect. Our findings underline the need for targeted smoking cessation support within specialist centers — especially for families where both parents smoke, those with financial or social difficulties, and for parents who have recently quit smoking to help prevent relapse. At the same time, stronger tobacco control policies are essential for Switzerland to make a smoke-free life easier for all. We need to find effective approaches to support parents of children with respiratory diseases when they visit the specialist center to quit smoking and maintain a smoke-free lifestyle.

## Data Availability

All data produced in the present study are available upon reasonable request to the authors.

## Acknowledgements

We thank all the families who took part in the SPAC study and the members of the SPAC Study Team. Members of the SPAC Study Team are: D. Mueller_-_Suter and P. Eng (Canton Hospital Aarau, Aarau, Switzerland); U. Frey, J. Hammer, A. Jochmann, D. Trachsel, and A. Oettlin (University Children’s Hospital Basel, Basel, Switzerland); P. Latzin, C. Abbas, M. Bullo, C. Casaulta, C. de Jong, E. Kieninger, I. Korten, L. Krüger, F. Singer, and S. Yammine (University Children’s Hospital Bern, Bern, Switzerland); P. Iseli (Children’s Hospital Chur, Chur, Switzerland); K. Hoyler (private paediatric pulmonologist, Horgen, Switzerland); S. Blanchon, S. Guerin, and I. Rochat (University Children’s Hospital Lausanne, Lausanne, Switzerland); N. Regamey, M. Lurà, M. Hitzler, K. Hrup, and J. Stritt (Children’s Hospital of Central Switzerland, Lucerne, Switzerland); J. Barben (Children’s Hospital St Gallen, St Gallen, Switzerland); O. Sutter (private pediatric practice, Worb, Bern, Switzerland); A. Moeller, A. Hector, K. Heschl, A. Jung, T. Schürmann, L. Thanikkel, and J. Usemann (University Children’s Hospital Zurich, Zurich, Switzerland); and M. Sasaki, M. Ganbat, R. Makhoul, B. Guerra, F. Romero, M. Goutaki and C. E. Kuehni.

We thank the Swiss National Cohort (SNC, www.swissnationalcohort.ch) for providing data on the Swiss-SEP index.

## Data availability statement

The data that support the findings of this study are available on reasonable request from the corresponding author.

## Ethical statement

The Bern Cantonal Ethics Committee (Kantonale Ethikkommission Bern 2016-02176) approved the study; written informed consent was obtained from parents and patients aged ≥ 14 years.

## List of abbreviations

SPAC: Swiss Paedatric Airway Cohort ETS Environmental Tobacco Smoke
IQR: Interquartile Range
CI: Confidence Interval
OR: Odds Ratio
Swiss-SEP: The Swiss Neighbourhood Index of Socioeconomic Position

## Supplementary items

**Figure S1.**
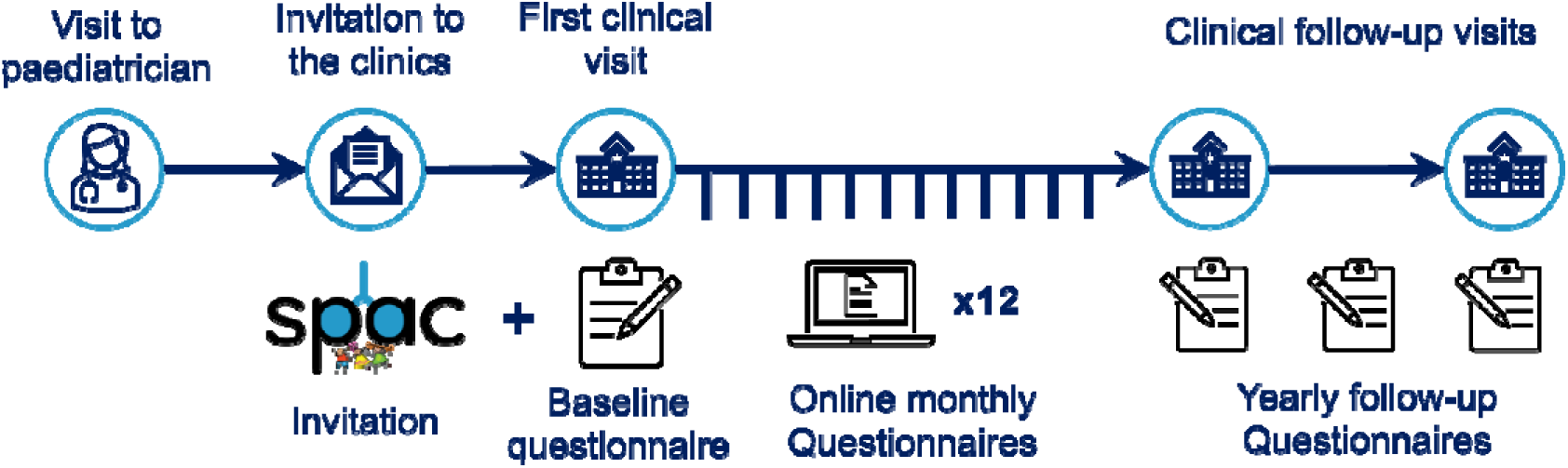
Swiss Paediatric Airway Cohort study design

**Figure S2.**
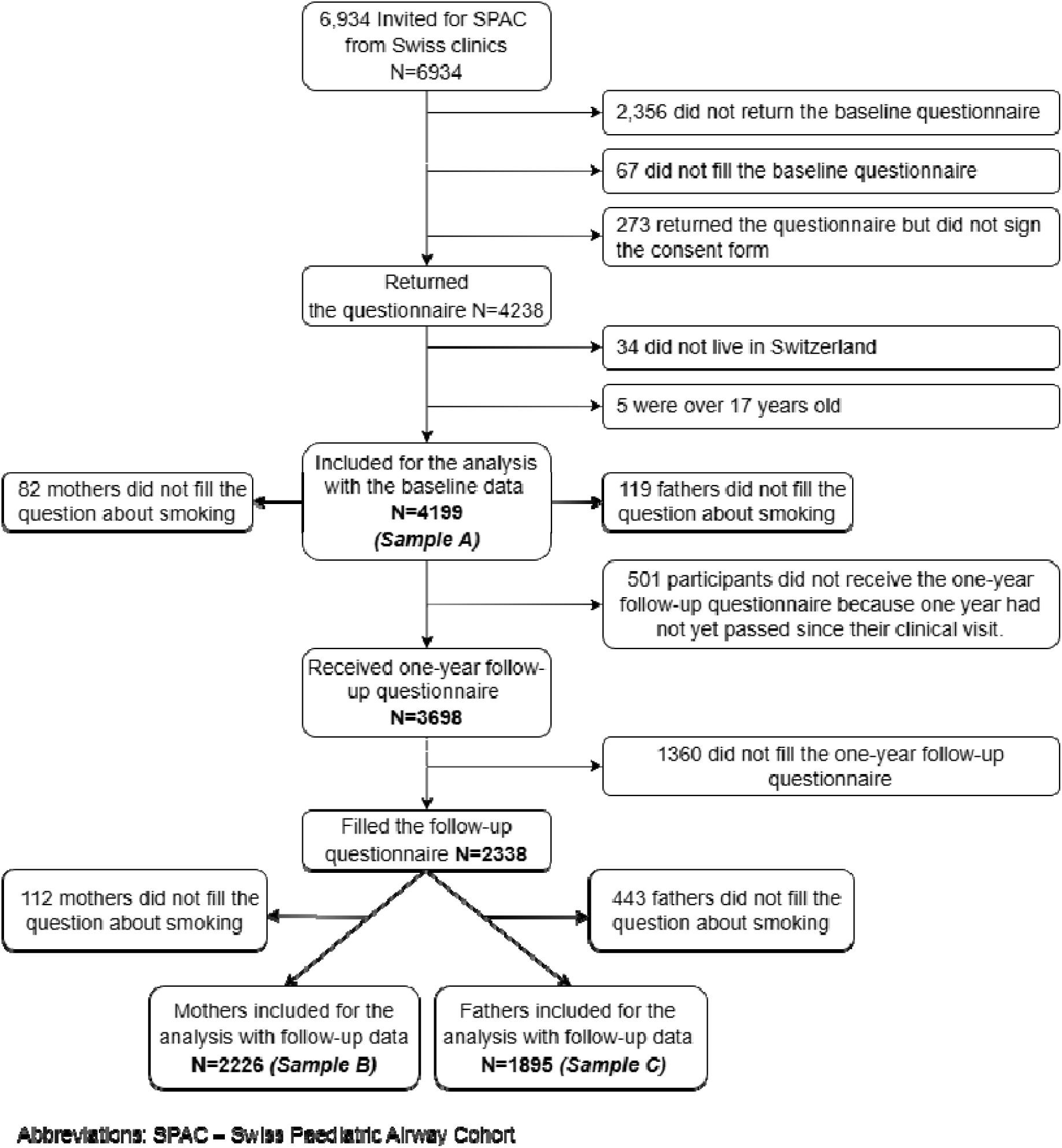
Flow diagram of the study population starting from children invited to SPAC to those included in the analysis. Sample A – used for the baseline analysis Sample B – used for assessing the changes in maternal smoking behavior over one year Sample C – used for assessing the changes in paternal smoking behavior over one year

**Table S1.**
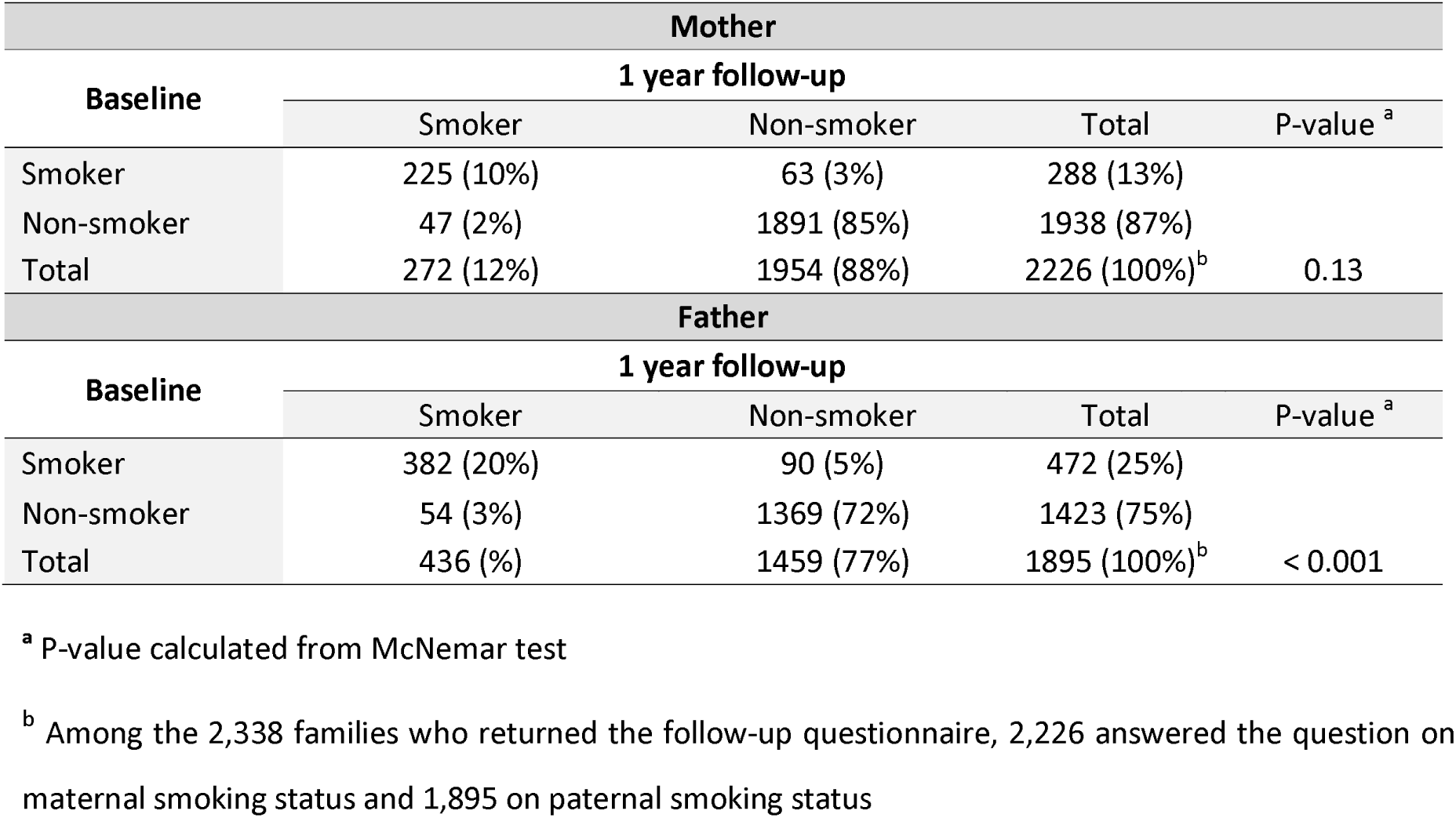
Changes in parental smoking status between initial clinical visit and one-year follow-up using McNemar test.

**Figure S3.**
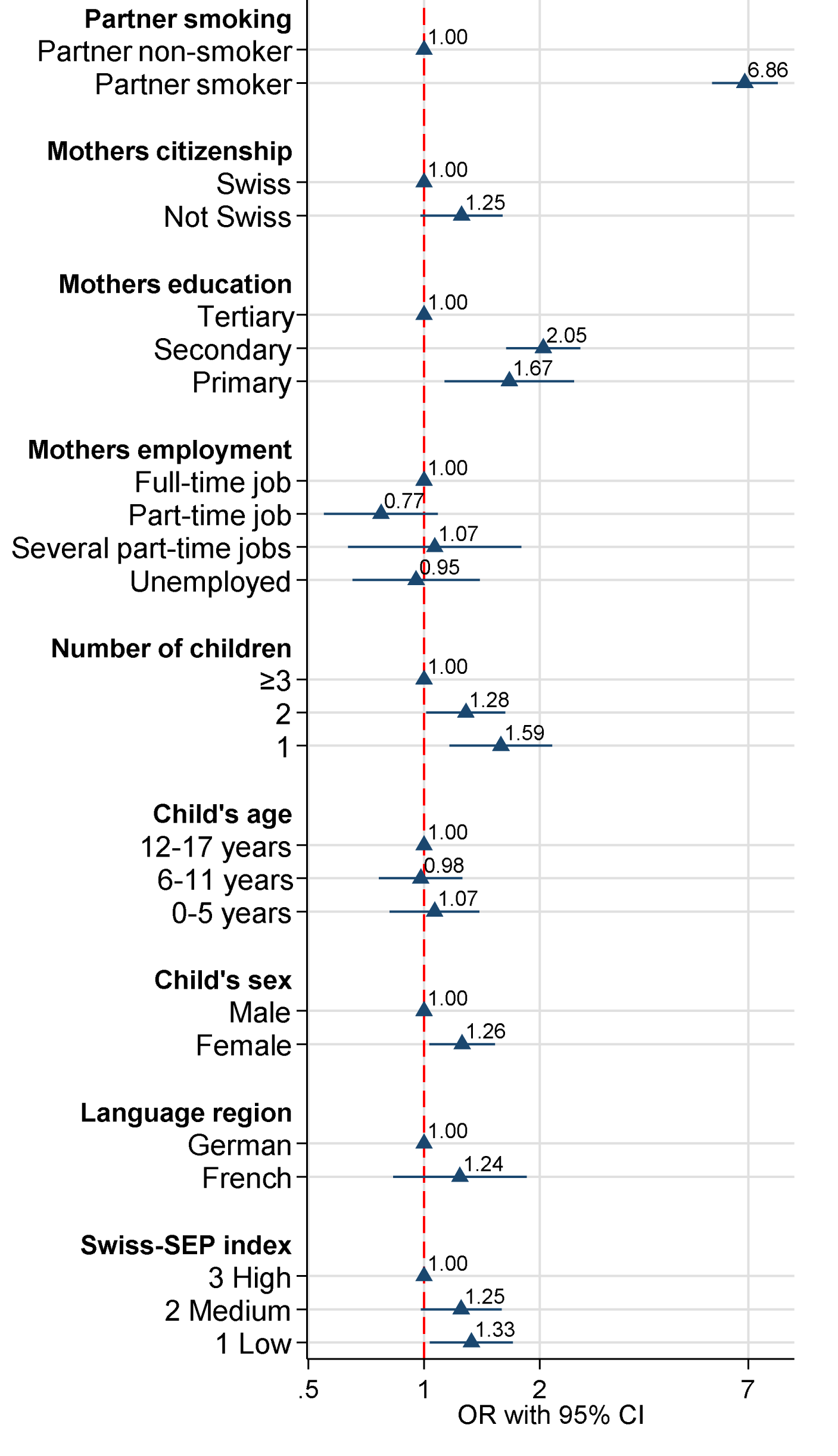
Predictors of maternal smoking at the baseline clinical visit in the Swiss Paediatric Airway Cohort, from multivariable logistic regression model (N=3759) Abbreviations: CI – confidence interval; OR – odds ratio; SPAC – Swiss Paediatric Airway Cohort; SEP – The Swiss Neighbourhood Index of Socioeconomic Position;

**Figure S4.**
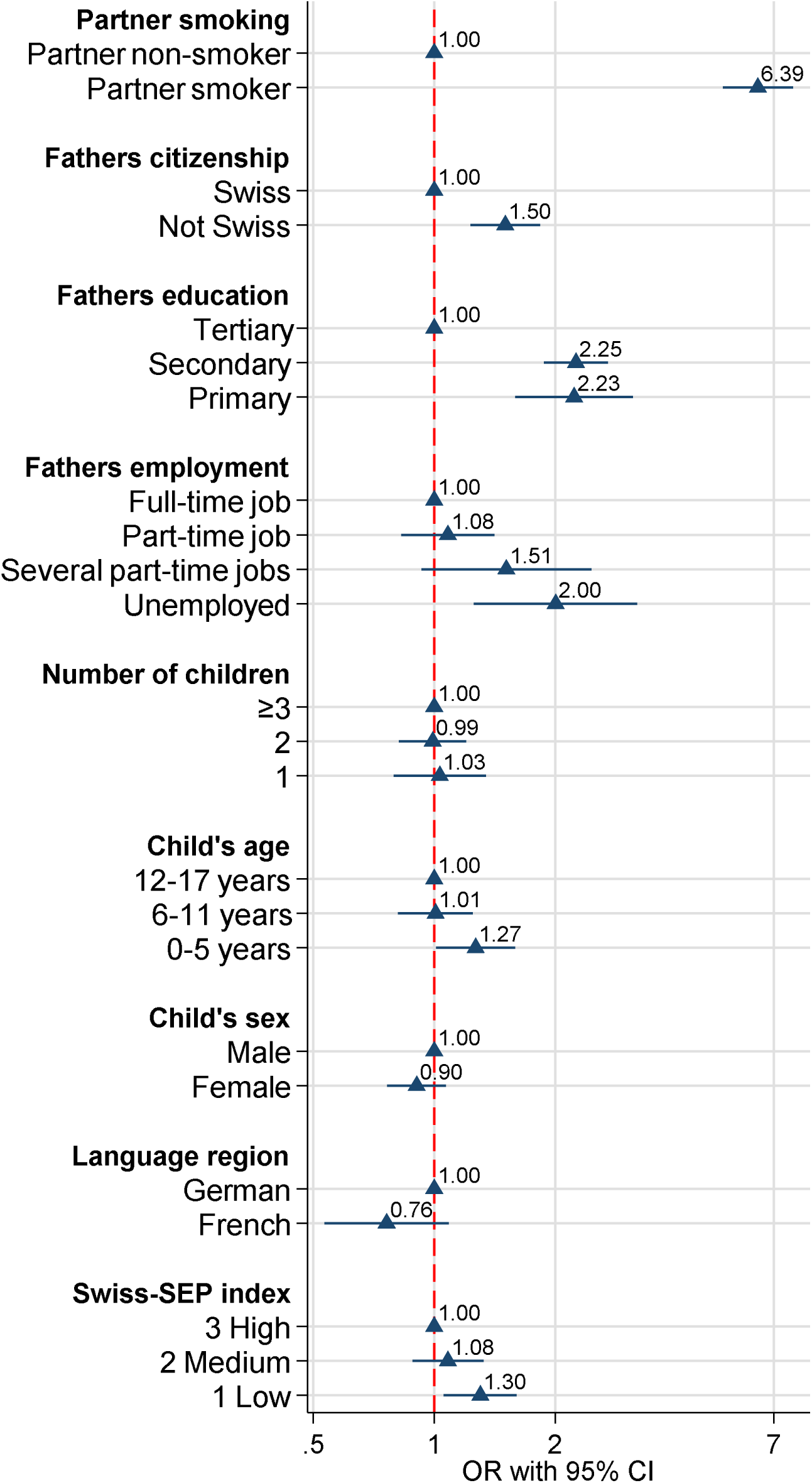
Predictors of paternal smoking at the baseline clinical visit in the Swiss Paediatric Airway Cohort, from multivariable logistic regression model (N=3696) Abbreviations: CI – confidence interval; OR – odds ratio; SPAC – Swiss Paediatric Airway Cohort; SEP – The Swiss Neighbourhood Index of Socioeconomic Position;

**Table S2.** Comparison of characteristics of responders and non-responders of the follow-up questionnaire among SPAC participants.

| Exposure |  | Non-responder<br>follow-up<br>N=1360 | Responder<br>follow-up<br>N=2338 | Total received follow-up questionnaire<br>N=3698 |
| --- | --- | --- | --- | --- |
| **Language region**<br>$P^b=0.874$ | German | 1,278 (94%) | 2,194 (94%) | 3,472 (94%) |
|  | French | 82 (6%) | 144 (6%) | 226 (6%) |
| **Child's age**<br>$P=0.008$ | 0-5 years | 428 (31%) | 743 (32%) | 1,171 (32%) |
|  | 6-11 years | 543 (40%) | 1,028 (44%) | 1,571 (42%) |
|  | 12-17 years | 389 (29%) | 567 (24%) | 956 (26%) |
| **Child's sex**<br>$P=0.342$ | Male | 798 (59%) | 1,409 (60%) | 2,207 (60%) |
|  | Female | 562 (41%) | 929 (40%) | 1,491 (40%) |
| **Number of children**<br>$P=0.664$ | 1 child | 220 (17%) | 360 (16%) | 580 (16%) |
|  | 2 children | 737 (56%) | 360 (56%) | 2,033 (56%) |
|  | ≥3 children | 362 (27%) | 652 (28%) | 1,014 (28%) |
| **Swiss-SEP index**<br>$P=0.506$ | 1 Low | 408 (30%) | 682 (29%) | 1,090 (29%) |
|  | 2 Medium | 461 (34%) | 768 (33%) | 1,229 (33%) |
|  | 3 High | 490 (36%) | 888 (38%) | 1,378 (37%) |
| **Household location**<br>$P=0.076$ | Urban | 845 (62%) | 1,366 (58%) | 2,211 (60%) |
|  | Periurban | 312 (23%) | 581 (25%) | 893 (24%) |
|  | Rural | 202 (15%) | 391 (17%) | 593 (16%) |
| **Mothers' citizenship**<br>$P<0.001$ | Swiss | 927 (71%) | 1,846 (80%) | 2,773 (77%) |
|  | Not Swiss | 385 (29%) | 460 (20%) | 845 (23%) |
| **Mothers' employment**<br>$P<0.001$ | Full-time job | 137 (11%) | 163 (7%) | 300 (8%) |
|  | Part-time job | 796 (62%) | 1,577 (69%) | 2,373 (66%) |
|  | Several part time jobs | 72 (6%) | 111 (5%) | 183 (5%) |
|  | Unemployed/education | 281 (22%) | 443 (19%) | 724 (20%) |
| **Mothers' education**<br>$P<0.001$ | Tertiary | 505 (39%) | 1,057 (46%) | 1,562 (43%) |
|  | Secondary | 646 (50%) | 1,110 (48%) | 1,756 (49%) |
|  | Primary | 146 (11%) | 131 (6%) | 277 (8%) |
| **Mothers' partner who smokes**<br>$P<0.001$ | Non-smoker | 877 (68%) | 1,720 (77%) | 2,597 (73%) |
|  | Smoker | 408 (32%) | 517 (23%) | 925 (26%) |
| **Fathers' citizenship**<br>$P<0.001$ | Swiss | 907 (70%) | 1,831 (80%) | 2,738 (76%) |
|  | Not Swiss | 387 (30%) | 455 (20%) | 842 (24%) |
| **Fathers' employment**<br>$P=0.042$ | Full-time job | 1,075 (86%) | 1,870 (83%) | 2,945 (84%) |
|  | Part-time job | 118 (9%) | 271 (12%) | 389 (11%) |
|  | Several part time jobs | 27 (2%) | 68 (3%) | 95 (3%) |
|  | Unemployed/education | 35 (3%) | 56 (2%) | 91 (3%) |
| **Fathers' education**<br>$P<0.001$ | Tertiary | 525 (41%) | 1,106 (49%) | 1,631 (46%) |
|  | Secondary | 625 (49%) | 1,027 (45%) | 1,652 (47%) |
|  | Primary | 127 (10%) | 138 (6%) | 265 (7%) |
| **Fathers' partner who smokes**<br>$P<0.001$ | Non-smoker | 1,039 (79%) | 2,014 (87%) | 3,053 (84%) |
|  | Smoker | 279 (21%) | 293 (13%) | 572 (16%) |

Abbreviations: SPAC – Swiss Paediatric Airway Cohort
<sup>b</sup> P is derived from the chi-squared test.

**Table S3.** Associations of maternal and paternal smoking with sociodemographic characteristics at the baseline clinical visit in the SPAC using cross tabulation and Pearson Chi2 test.

| Exposure |  | Mother at baseline |  |  | Father at baseline |  |  |
| --- | --- | --- | --- | --- | --- | --- | --- |
|  |  | Non-Smoker | Smoker | Total | Non-smoker | Smoker | Total |
|  |  | N=3478 | N=639 | N=4199 | N=2955 | N=1041 | N=4199 |
| Language region | German | 3,266 (85%) | 596 (15%) | 3,862 | 2,766 (74%) | 989 (26%) | 3,755 |
|  | French | 157 (82%) | 34 (18%) | 191 | 135 (75%) | 46 (25%) | 181 |
|  |  | P<sup>a</sup>=0.378 |  |  | P<sup>a</sup>=0.783 |  |  |
| Child's age | 0-5 years | 1,074 (83%) | 225 (17%) | 1,299 | 898 (70%) | 376 (31%) | 1,274 |
|  | 6-11 years | 1,526 (85%) | 263 (15%) | 1,789 | 1,313 (76%) | 424 (24%) | 1,737 |
|  | 12-17 years | 878 (85%) | 151 (15%) | 1,029 | 744 (76%) | 241 (24%) | 985 |
|  |  | P= 0.096 |  |  | **P=0.003** |  |  |
| Child's sex | Male | 2,101 (85%) | 366 (15%) | 2,467 | 1,767 (74%) | 631 (26%) | 2,398 |
|  | Female | 1,377 (83%) | 273 (17%) | 1,650 | 1,188 (74%) | 410 (26%) | 1,598 |
|  |  | P=0.138 |  |  | P=0.643 |  |  |
| Number of children | 1 child | 526 (81%) | 123 (19%) | 649 | 438 (70%) | 186 (30%) | 624 |
|  | 2 children | 1,937 (85%) | 353 (15%) | 2,290 | 1,665 (75%) | 563 (25%) | 2,228 |
|  | ≥3 children | 973 (87%) | 150 (13%) | 1,123 | 812 (75%) | 276 (25%) | 1,088 |
|  |  | **P=0.007** |  |  | P=0.063 |  |  |
| Education | Tertiary | 1,637 (91%) | 165 (9%) | 1,802 | 1,528 (84%) | 285 (16%) | 1,813 |
|  | Secondary | 1,540 (80%) | 386 (20%) | 1,926 | 1,188 (66%) | 606 (34%) | 1,794 |
|  | Primary | 223 (76%) | 69 (24%) | 292 | 160 (60%) | 108 (40%) | 268 |
|  |  | **P<0.001** |  |  | **P<0.001** |  |  |
| Citizenship | Swiss | 2,640 (85%) | 452 (15%) | 3,092 | 2,291 (77%) | 699 (23%) | 2,990 |
|  | Not Swiss | 785 (82%) | 175 (18%) | 960 | 605 (66%) | 318 (34%) | 923 |
|  |  | **P=0.007** |  |  | **P<0.001** |  |  |
| Employment | Full-time job | 274 (79%) | 74 (21%) | 348 | 2,389 (74%) | 821 (26%) | 3,210 |
|  | Part-time job | 2,299 (86%) | 371 (14%) | 2,670 | 348 (78%) | 98(22%) | 446 |
|  | Several part time jobs | 167 (83%) | 34 (17%) | 201 | 69 (69%) | 31 (31%) | 100 |
|  | Unemployed, education | 657 (83%) | 134 (17%) | 791 | 56 (57%) | 42 (43%) | 98 |
|  |  | **P=0.001** |  |  | **P<0.001** |  |  |
| Swiss-SEP index | 1 Low | 947 (79%) | 251 (21%) | 1,198 | 774 (66%) | 392 (34%) | 1,166 |
|  | 2 Medium | 1,140 (84%) | 219 (16%) | 1,359 | 977 (74%) | 345 (26%) | 1,322 |
|  | 3 High | 1,390 (89%) | 169 (11%) | 1,559 | 1,203 (80%) | 304 (20%) | 1,507 |
|  |  | **P<0.001** |  |  | **P<0.001** |  |  |
| Household location | Urban | 2,091 (85%) | 379 (15%) | 2,470 | 1,769 (74%) | 623 (26%) | 2,392 |
|  | Periurban | 837 (85%) | 153 (15%) | 990 | 729 (76%) | 233 (24%) | 962 |
|  | Rural | 549 (84%) | 107 (16%) | 656 | 456 (71%) | 185 (29%) | 641 |
|  |  | P=0.829 |  |  | P=0.116 |  |  |
| Partner who smokes | Non-smoker | 2,724 (81%) | 620 (19%) | 3,344 | 2,724 (92%) | 225 (8%) | 2,949 |
|  | Smoker | 225 (37%) | 390 (63%) | 615 | 620 (61%) | 390 (39%) | 1,010 |
|  |  | **P<0.001** |  |  | **P<0.001** |  |  |

Abbreviations: SPAC – Swiss Paediatric Airway Cohort
<sup>a</sup>P is derived from the chi-squared test.

